# Acute Endotheliitis (Type 3 Hypersensitivity Vasculitis) in Ten COVID-19 Autopsy Brains

**DOI:** 10.1101/2021.01.16.21249632

**Authors:** Roy H. Rhodes, Gordon L. Love, Fernanda Da Silva Lameira, Maryam Shahmirzadi Sadough, Sharon E. Fox, Richard S. Vander Heide

## Abstract

Central nervous system (CNS) involvement in COVID-19 may occur through direct SARS-CoV-2 invasion through peripheral or cranial nerves or through vascular endothelial cell infection. The renin-angiotensin system may play a major part in CNS morbidity. Effects of hypoxia have also been implicated in CNS lesions in COVID-19. This communication reports on ten consecutive autopsies of individuals with death due to COVID-19 with decedent survival ranging from 30 minutes to 84 days after admission. All ten brains examined had neutrophilic microvascular endotheliitis present in variable amounts and variably distributed. Importantly, this acute stage of type 3 hypersensitivity vasculitis can be followed by fibrinoid necrosis and inner vascular wall sclerosis, but these later stages were not found. These results suggest that a vasculitis with autoimmune features occurred in all ten patients. It is possible that viral antigen in or on microvascular walls or other antigen-antibody complexes occurred in all ten patients proximate to death as a form of autoimmune vasculitis.

## Introduction

The SARS-CoV-2 pandemic of 2020 involves injury to the major organs including the central nervous system (CNS). The mode of CNS involvement has been under consideration almost since the disease caused by SARS-CoV-2 (COVID-19) was reported. The main proposed modes of CNS involvement include direct olfactory nerve or cranial nerve infection with trans-synaptic CNS entry into olfactory bulbs and the ventrolateral medulla, respectively;^1-5^ a hematogenous route with CNS vascular endothelial cell infection allowing direct viral entry into the brain; and indirect CNS damage through a coagulopathy complicated by the formation of microthrombi or major vessel obstructive thrombosis.^1,2,6,7^ This indirect CNS damage may occur through a virus-induced dysfunction or imbalance of the renin-angiotensin system (RAS).^4,8,9^ Autoimmunity may play a role in CNS damage with the development of hypercytokinemia (“cytokine storm”) and type 3 hypersensitivity vasculitis.^1,2,10-17^ In addition, hypoxia in these patients affects the CNS as a result of a respiratory infection and coagulopathy found in most hospitalized patients.^1,2,6,18,19^

Cellular inflammation has been reported as a less prominent component of CNS damage in COVID-19. Vascular wall inflammation in a few autopsy reports consists of lymphocytes^18,20^ or mild focal chronic perivasculitis or vasculitis.^21-23^ Acute endotheliitis, the early component of type 3 hypersensitivity vasculitis, has not been demonstrated to date.^18,21^

COVID-19 autopsies early in the pandemic allowed the study of most major organs in conjunction with hospital engineering assistance to assure safe conditions in the autopsy room for the personnel involved, including by our group.^24^ Once our appropriate calvaria removal saw was obtained, the brain could also be examined.

The present report highlights microvascular findings, particularly acute neutrophilic endotheliitis, that may be relevant to neurological symptoms and perhaps death in COVID-19. These microvascular alterations were seen in the first ten consecutive autopsy brains available for study.

## Brief Clinical Summary of the Cohort

The ten autopsy cases included adults from middle age to elderly, half being female, with seven African Americans in the cohort. Survival after arrival at the hospital was under an hour to over 80 days (mean survival 19.93 ± 26.2 days). All patients had a confirmed RT-PCR–positive SARS-CoV-2 test and clinical findings typical of COVID-19 as previously described in our region’s population. Comorbidities in this cohort, particularly hypertension, were also typical for this population.^24^ Respiratory distress or acute respiratory failure at admission were common findings in this cohort. A review of times of death revealed no deaths close to the 7 AM hour when failure of the switch from automatic to voluntary breathing occurs. Table 1 lists clinical findings in these cases, with the patients placed in the ascending order of survival. Two patients had no past medical history available. These studies were determined to be exempt by the IRB at Louisiana State University Health Sciences Center, New Orleans.

**Table 1.**
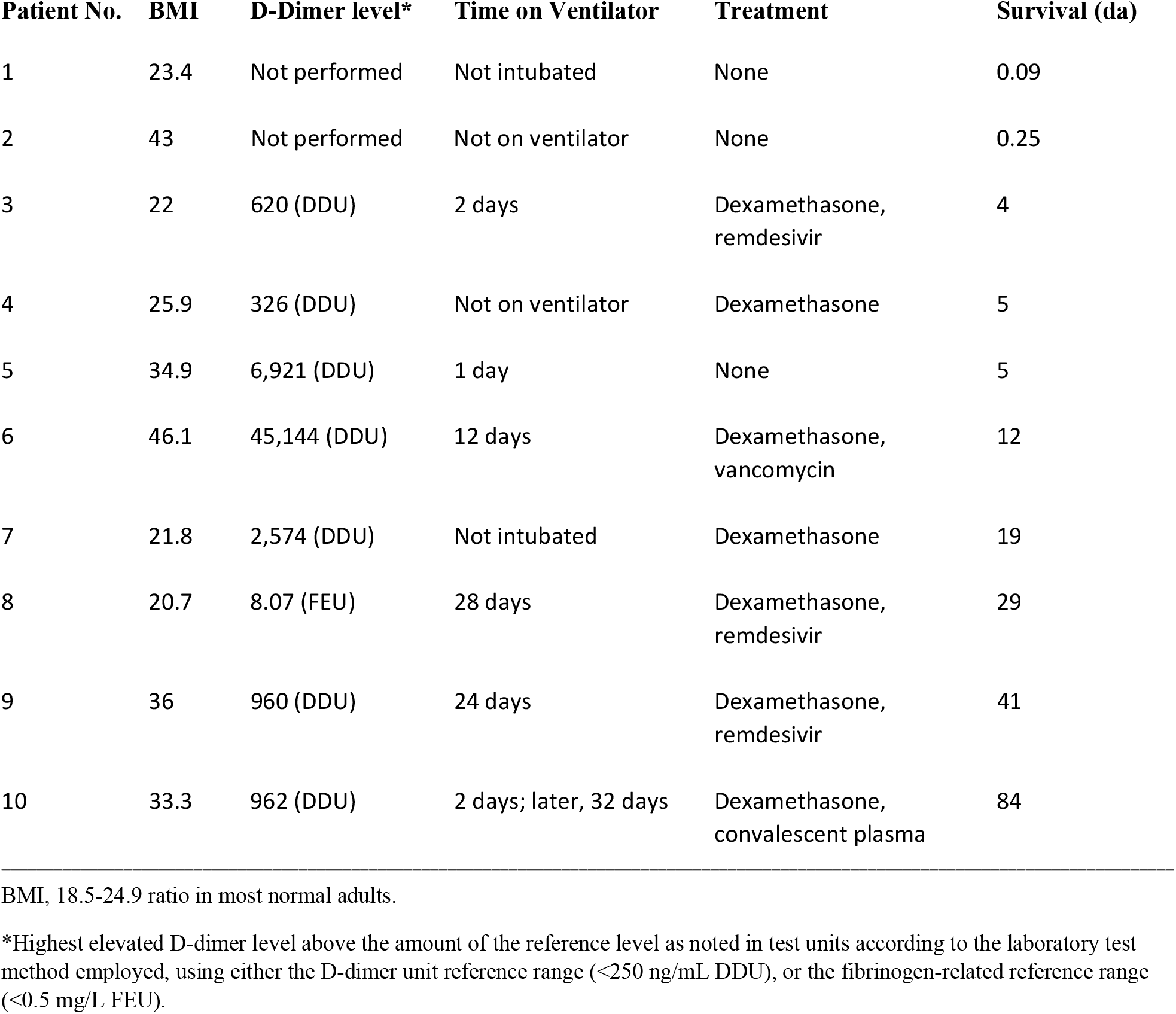
Clinical Findings of Ten COVID-19 Autopsy Patients.

## Autopsy Brain Findings

Table 2 lists relevant gross and histopathologic brain findings. Histopathologic examination included between 32 and 50 paraffin-embedded tissue blocks for each brain. Patient 1, who was pronounced dead soon after arrival at the Emergency Department, suffered large hemorrhages and associated recent cerebral infarcts with significant infiltration of polymorphonuclear neutrophils (PMNs). Similar lesions were present in the midbrain and pons, while acute encephalitis was present in the medulla. Acute and chronic leptomeningitis was found, and acute arteritis was identified in the vertebral artery in the posterior fossa and in two small leptomeningeal arteries. Bacterial and fungal stains did not demonstrate organisms and Congo Red stain with polarization revealed no vascular-wall amyloid. For patient 5, bilateral cerebellar tonsillar herniations were associated with global hypoxic nerve-cell change. Patient 8 had thrombotic obstruction of the left common carotid artery and an atherosclerotic plaque in the right common carotid artery in the neck demonstrated by radiologic imaging with large bilateral intermediate cerebral infarcts resulting. In the other cases, gross findings were typical for age and comorbidities. Most cases had histopathologic evidence of microvascular mural adventitial collagenous thickening. Although two cases had no available past history, the latter finding supports frequent microvascular injury from systemic hypertension in the cohort.

**Table 2.**
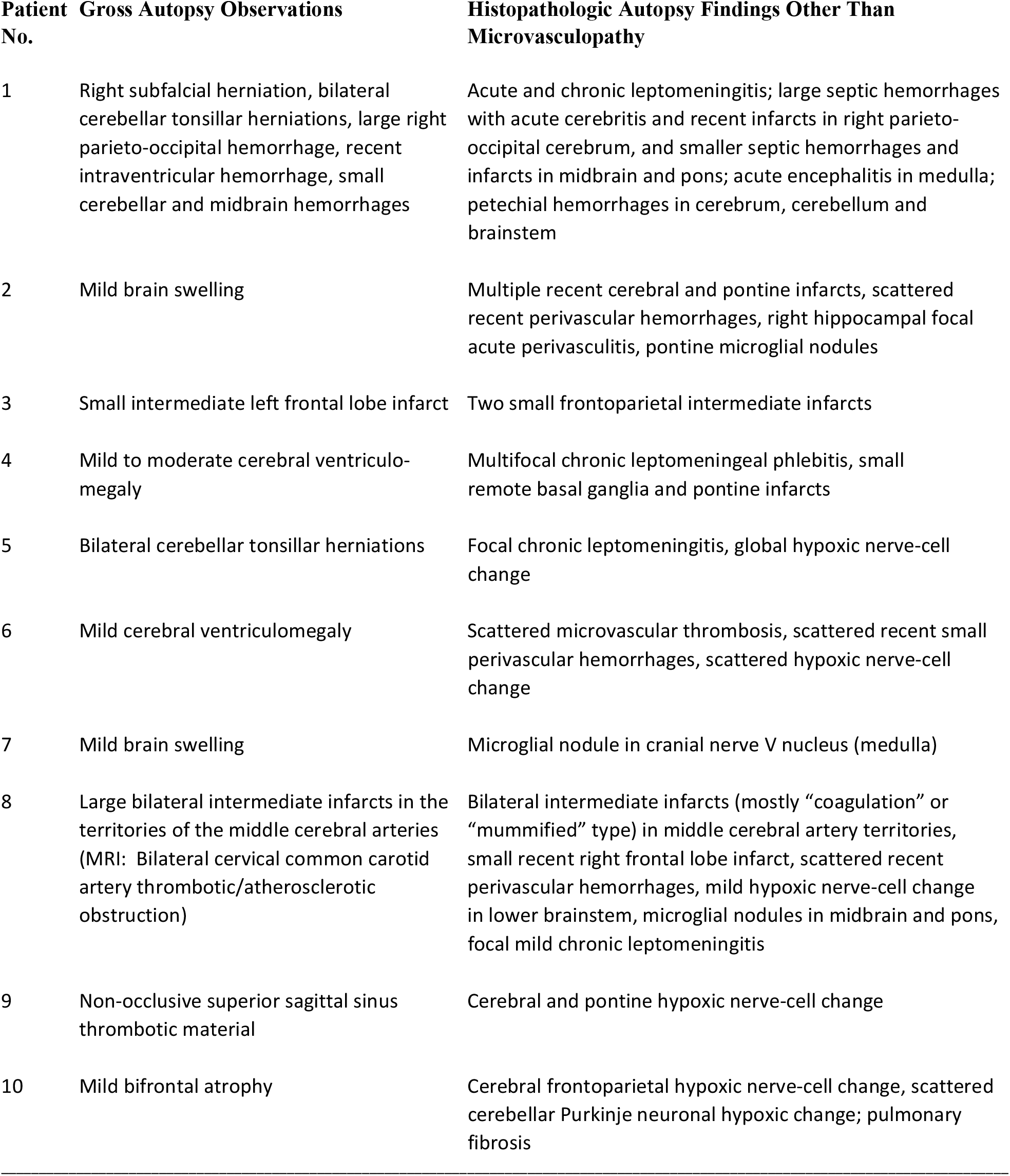
Autopsy Findings for Ten COVID-19 Patients.

Not all cases had hypoxic change or infarcts, and few cases had petechial hemorrhages. Small, scattered perivascular hemorrhages around microvessels were frequently found. All ten cases had acute neutrophilic endotheliitis with a variable amount and distribution in each case. The recognition of acute neutrophilic endotheliitis was made in microvessels with intraluminal PMNs that had apoptotic bodies (nuclear dust, or “leukocytoclasis”). There were a few dehiscent microvascular walls that may have represented mural necrosis, although some of these were largely obscured by blood cells. Five cases had acute perivasculitis adjacent to one or a few microvessels. The full distribution of these microvascular findings appears in Table 3.

**Table 3.** Anatomic Sites of Acute Neutrophilic Endotheliitis, Perivasculitis and Vasculitis in Brain*.

| **Patients** | **Microvascular Acute Neutrophilic Endotheliitis** | **Acute Perivasculitis and Arteritis** |
| --- | --- | --- |
| 1 | Caudal left midbrain (crus cerebri), rostral right basis pontis | Acute perivasculitis, rostral left midbrain tegmentum; acute arteritis, small subarachnoid artery walls adjacent to acute leptomeningitis |
| 2 | Occipital white matter, putamen, bilateral thalamus, anterior perforated substance (basal forebrain olfactory area), cerebellar lateral lobes, multiple levels of midbrain and pontine tegmentum (including locus coeruleus), multiple levels of basis pontis, multiple levels of tegmentum in medulla | Acute perivasculitis, right medial temporal lobe white matter and hippocampus |
| 3 | Left frontal white matter, within an intermediate infarct in left frontal lobe, white matter lateral to the tail of the left caudate nucleus, right hippocampus, right lateral lobe of the cerebellum |  |
| 4 | Right frontal pole white matter, left inferomedial temporal archicortex, right lateral temporal lobe white matter, right parietal cortex, right calcarine cortex, right thalamus and putamen, internal capsule, left superior colliculus, right midbrain tectum, caudal midbrain tegmentum, rostral basis pontis, medulla near inferior olivary nucleus | Acute perivasculitis, right substantia nigra and caudal midbrain tegmentum |
| 5 | Right lateral temporal lobe cortex and white matter, right internal capsule, cerebellar white matter, right pontine tegmentum, left pontine locus coeruleus and basis pontis (several levels) | Acute perivasculitis, left cerebellar folial white matter |
| 6 | Bilateral frontal pole and parietal cortex, right temporal presubicular white matter, left lateral temporal cortex, right occipital cortex, left occipital white matter, right putamen, bilateral thalamus, left basal forebrain, right lateral lobe of cerebellum, rostral right midbrain, caudal crus cerebri and substantia nigra, basis pontis | Acute perivasculitis, rostral left basis pontis |
| 7 | Right mid basis pontis |  |
| 8 | Caudal pontine tegmentum |  |
| 9 | Right frontal pole cortex, left temporal and parietal cortex, left thalamus, left putamen, bilateral globus pallidus externa, white matter of lateral cerebellar lobes, right rostral and caudal midbrain tegmentum and substantia nigra, left rostral substantia nigra, right caudal crus cerebri, right rostral basis pontis, pyramidal tract |  |
| 10 | Temporal cortex, hippocampi, thalamus, left internal capsule, cerebellum, pontine tegmentum, basis pontis, medulla reticular formation near nucleus/tractus solitarius (which had two thrombosed microvessels) |  |

\*See text for description of the phases of type 3 hypersensitivity vasculitis; no fibrinoid necrosis or findings of sclerosing intimal vasculitis were found.

Representative microvascular samples of acute endotheliitis and acute perivasculitis are shown in Figures 1–12.

**Fig. 1.**
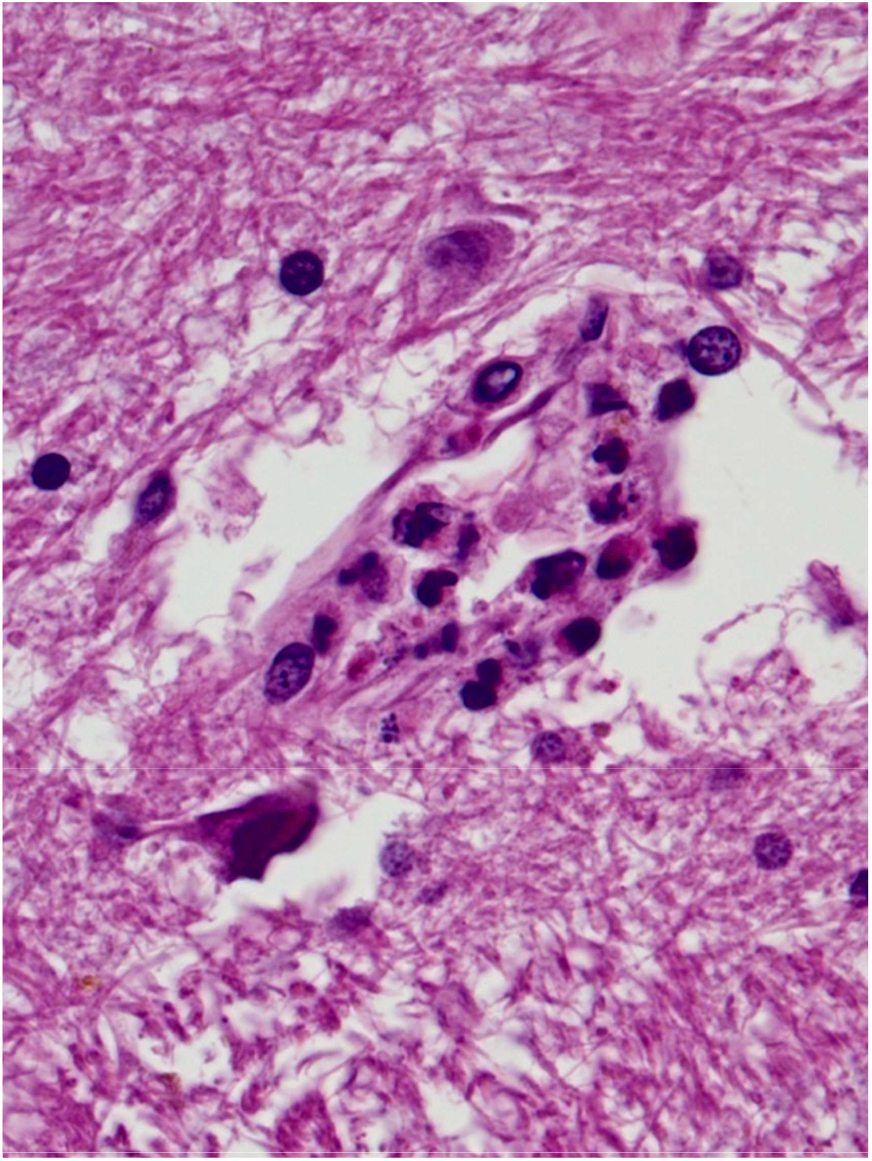
Patient 1: A microvessel in a recent infarct of the rostral right basis pontis has acute endotheliitis shown by leukocytoclasis (“nuclear dust”), or apoptosis, and some PMNs have transmigrated from the lumen to the microvascular adventitia, x400. H&E stain.

**Fig. 2.**
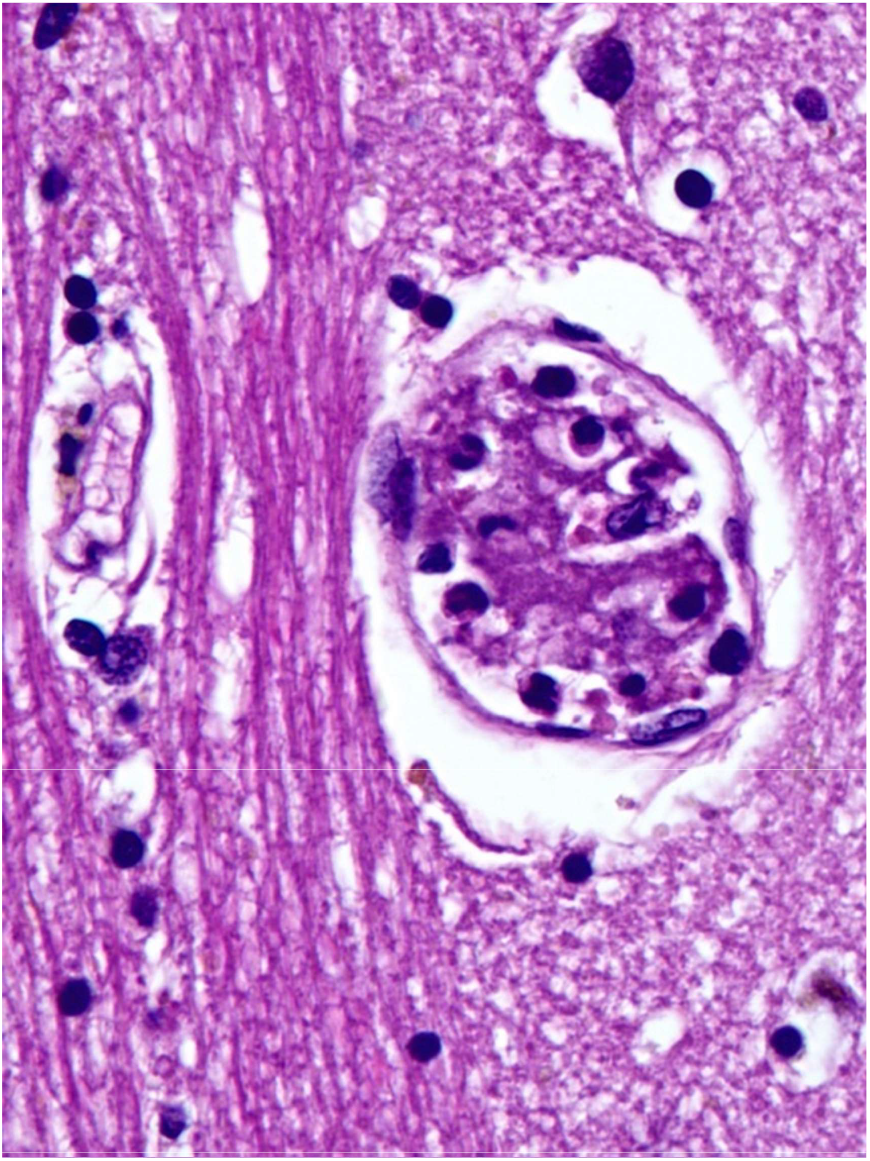
Patient 2: A microvessel in the left putamen has acute endo-theliitis and thrombosis, x400. H&E stain.

**Fig. 3.**
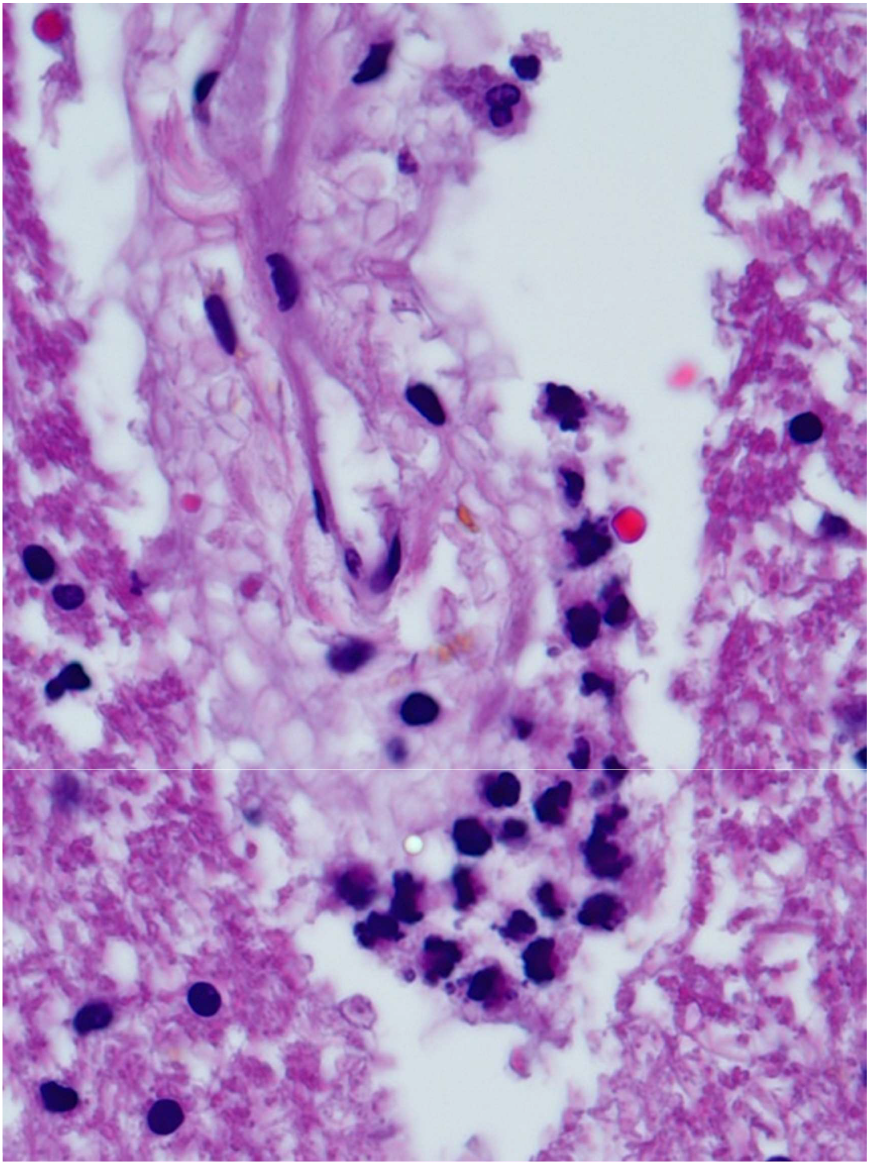
Patient 2: A microvessel in the right inferior temporal fusiform gyrus white matter has acute (“leukocytoclastic”) perivasculitis, x400. H&E stain.

**Fig. 4.**
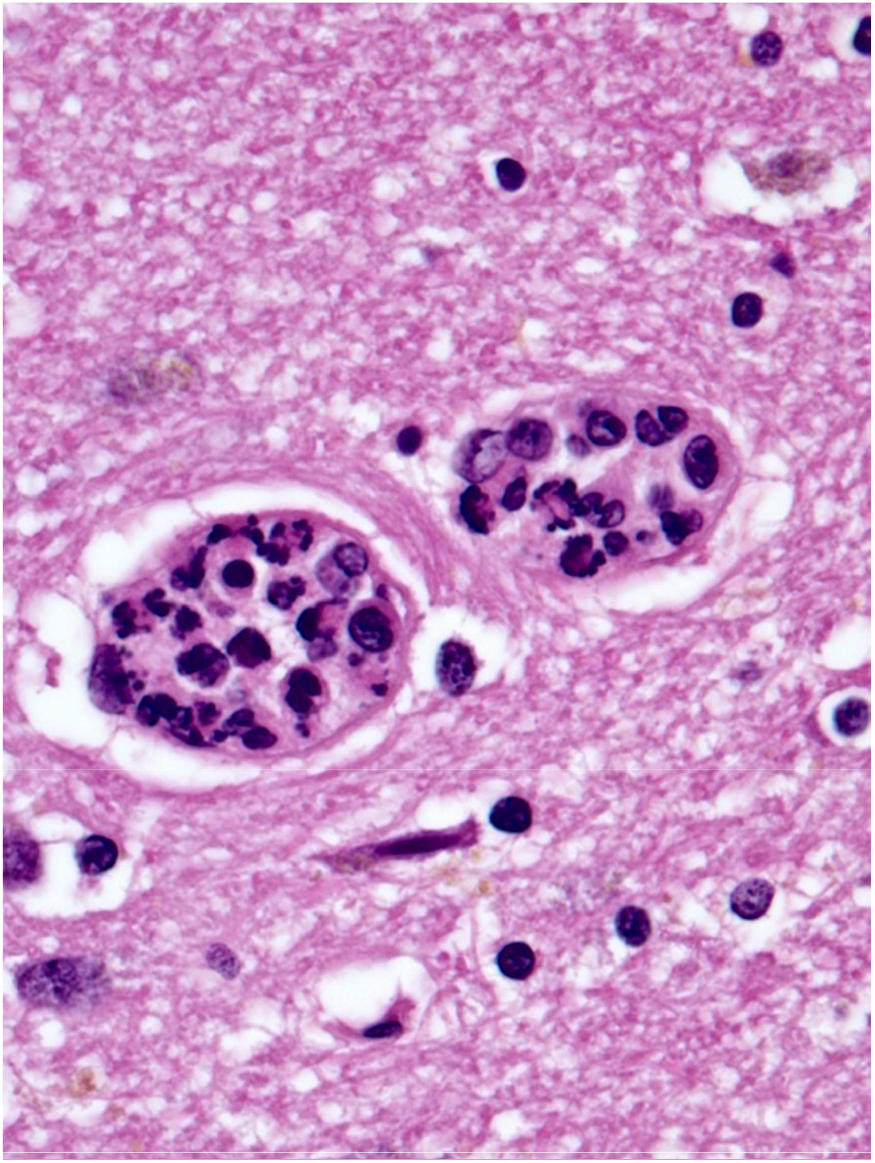
Patient 4: Microvessels in the right temporal lobe white matter have acute endotheliitis, x400. H&E stain.

**Fig. 5.**
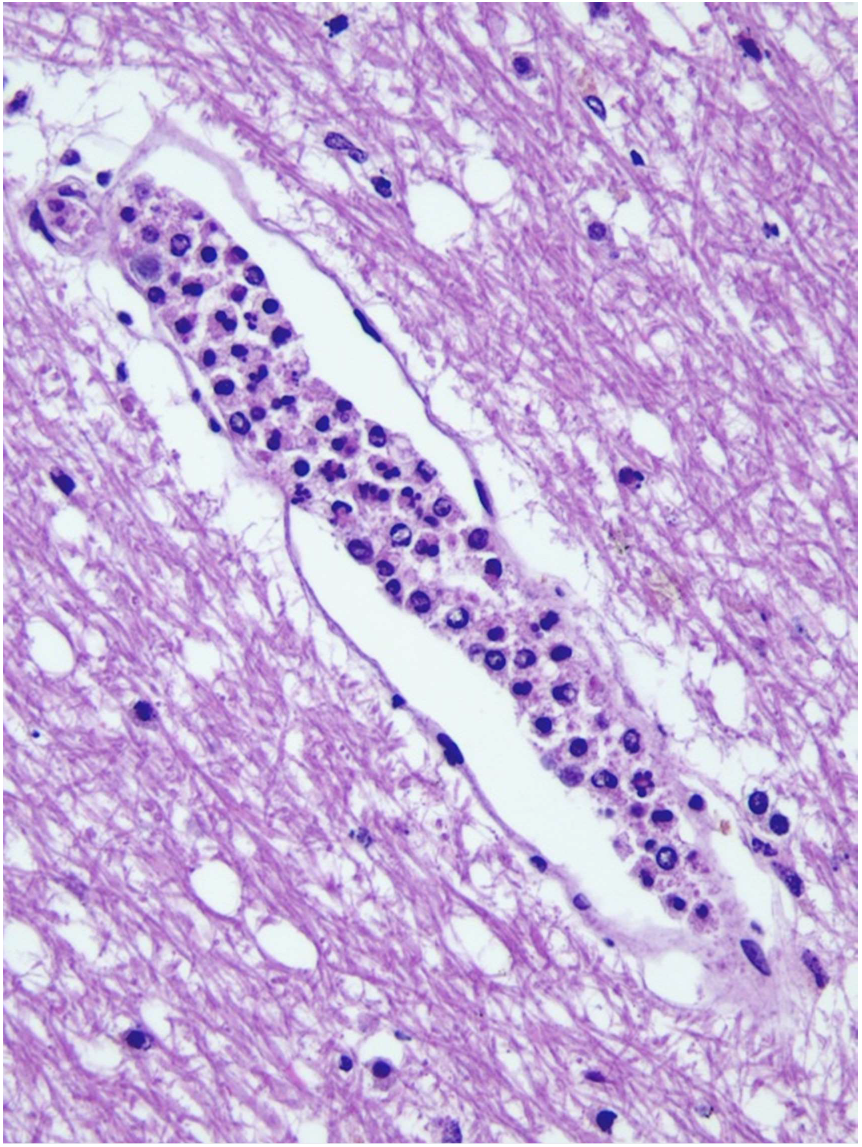
Patient 5: Dilated microvessel in the left cerebellar deep white matter has acute endotheliitis, x200. H&E stain.

**Fig. 6.**
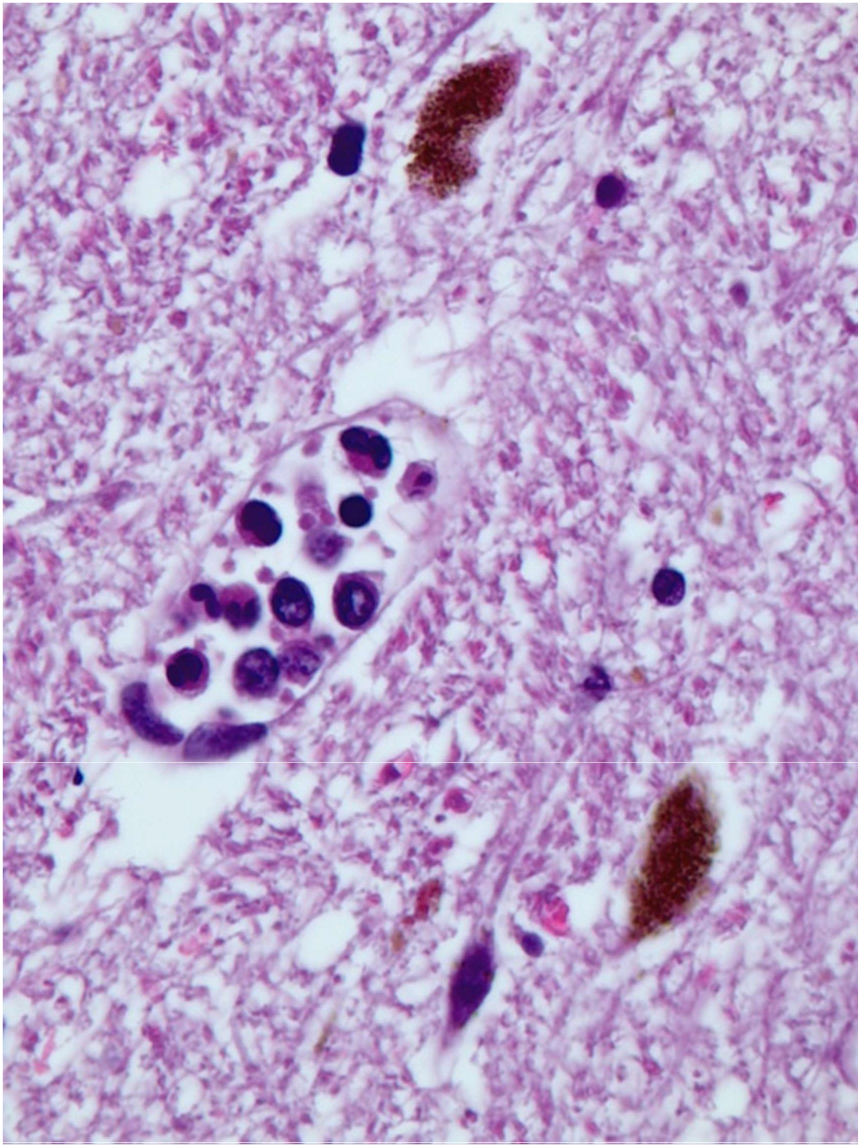
Patient 5: Dilated microvessel in the locus coeruleus of the left mid-level pontine tegmentum has acute endotheliitis, x400. H&E stain.

**Fig. 7.**
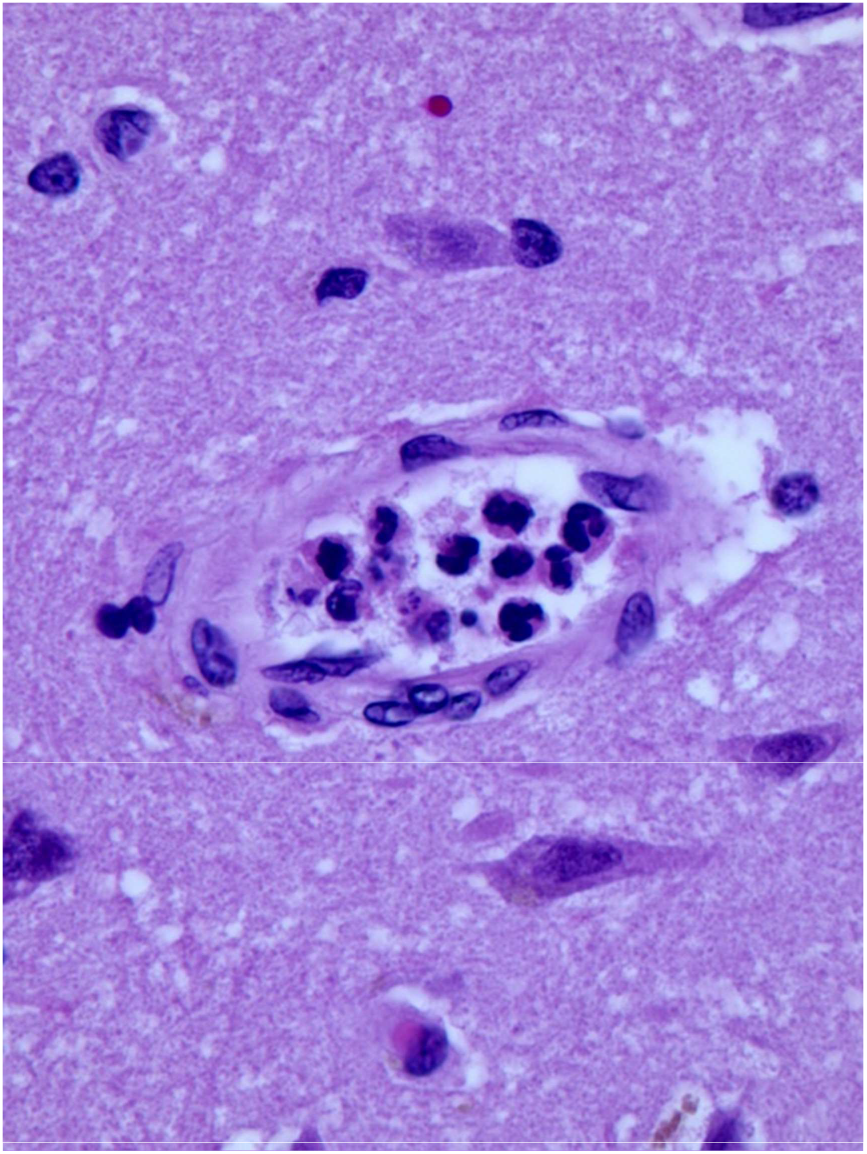
Patient 6: Microvessel in the left parietal boundary-zone neocortex has acute endotheliitis, x400. H&E stain.

**Fig. 8.**
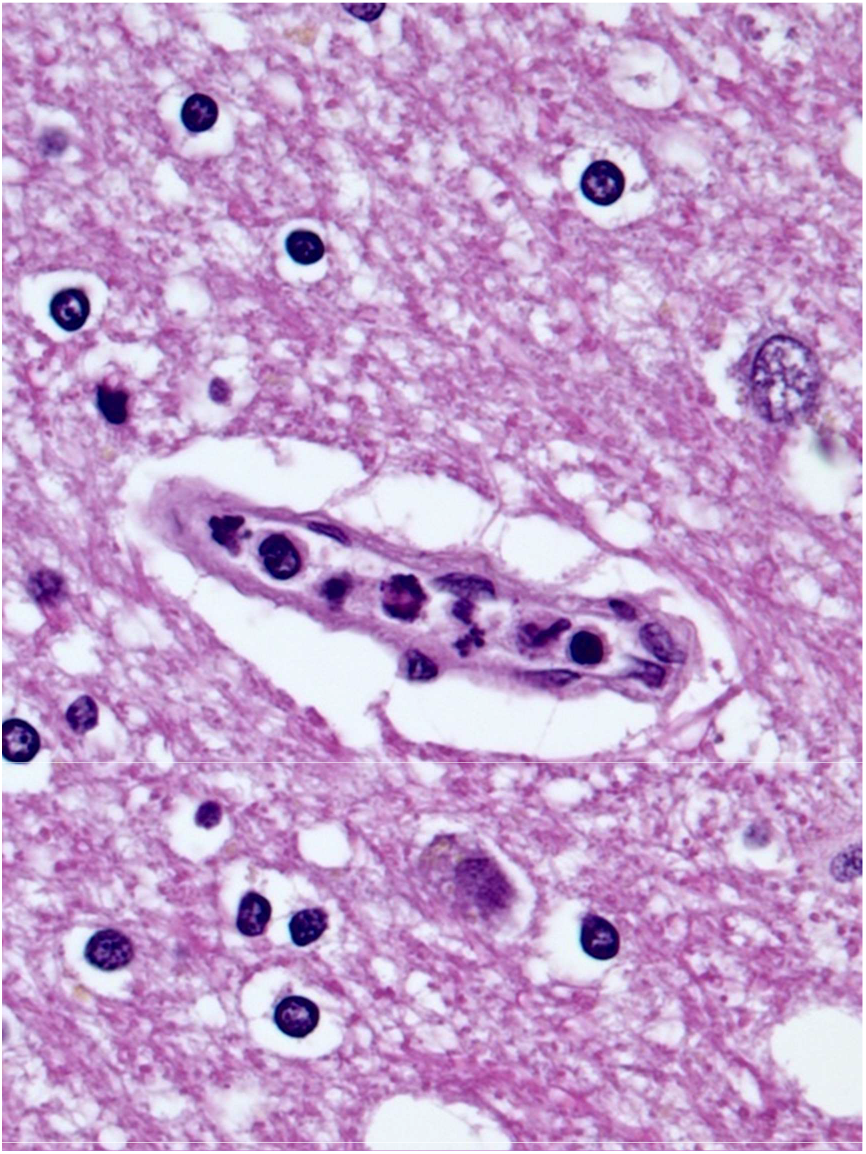
Patient 6: Capillary with acute endotheliitis in the left internal capsule, x400. H&E stain.

**Fig. 9.**
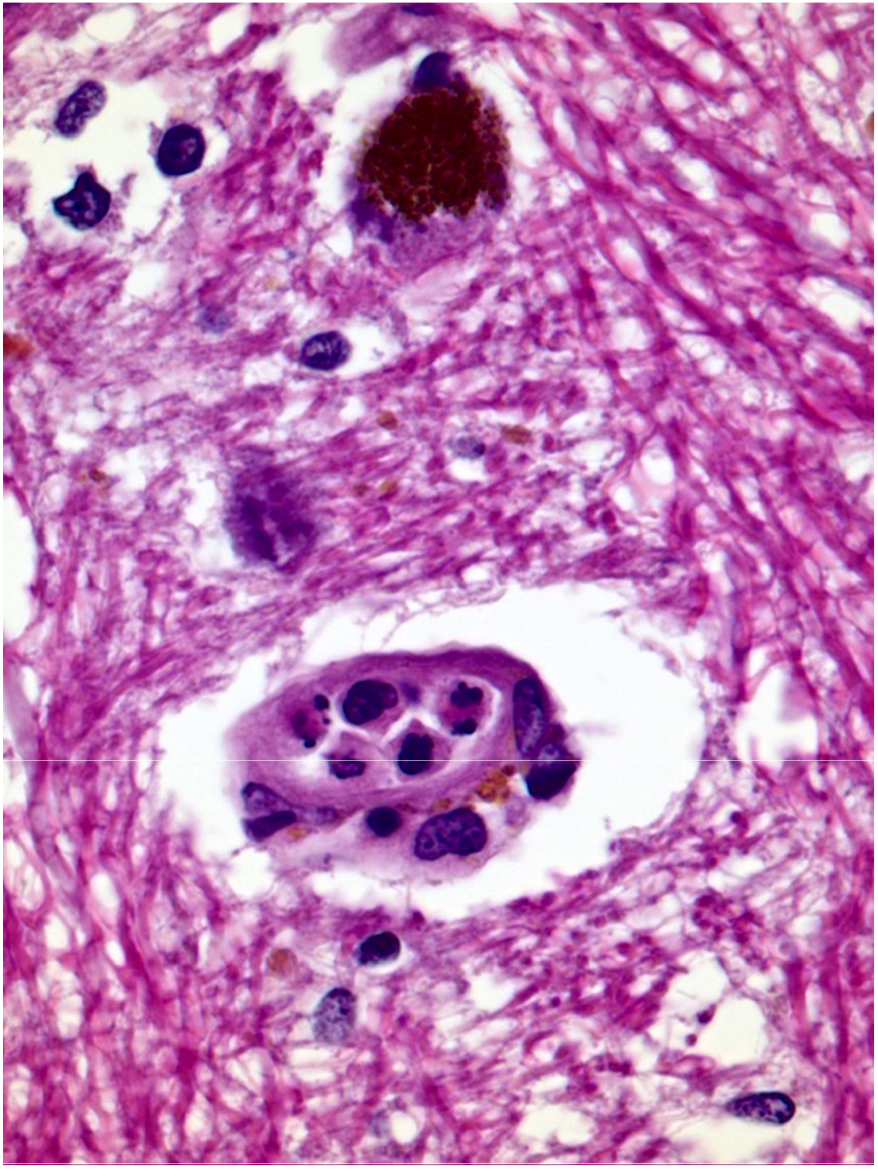
Patient 6: Microvessel with acute endotheliitis in the substantia nigra of the caudal midbrain, x400. H&E stain.

**Fig. 10.**
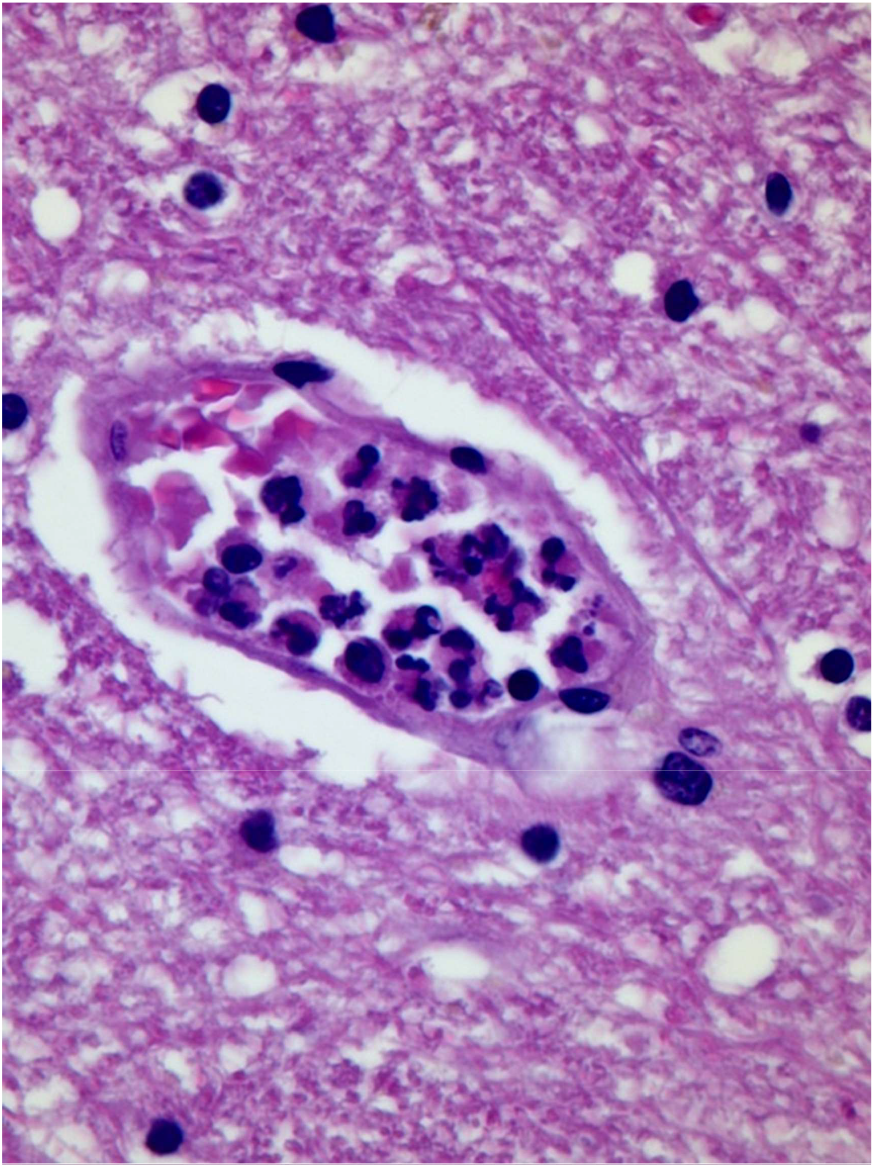
Patient 9: Microvessel in the left thalamus with acute endotheliitis, x400. H&E stain.

**Fig. 11.**
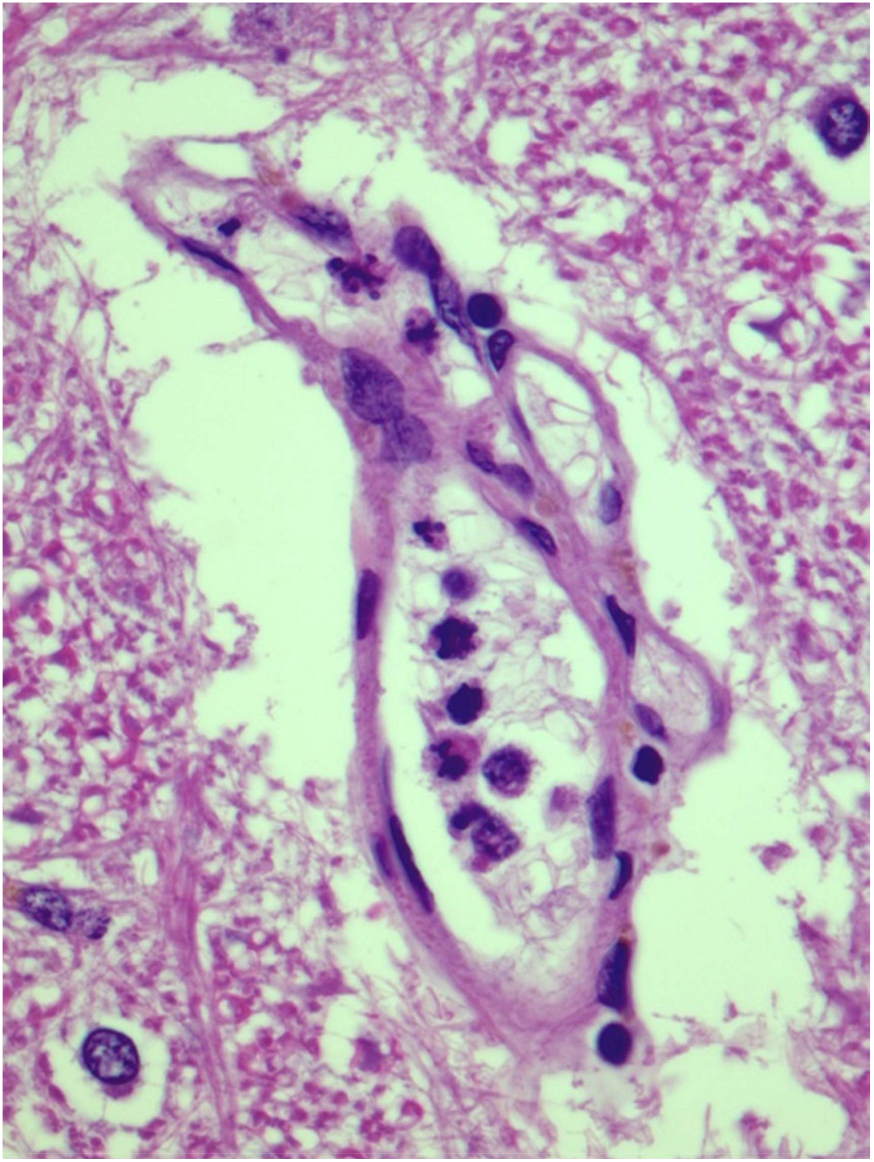
Patient 10: A microvessel in the dorsomedial reticular formation in the mid-level medulla has acute endotheliitis. The microvessel appears to be undergoing intussusceptive arborization, x400. H&E stain.

**Fig. 12.**
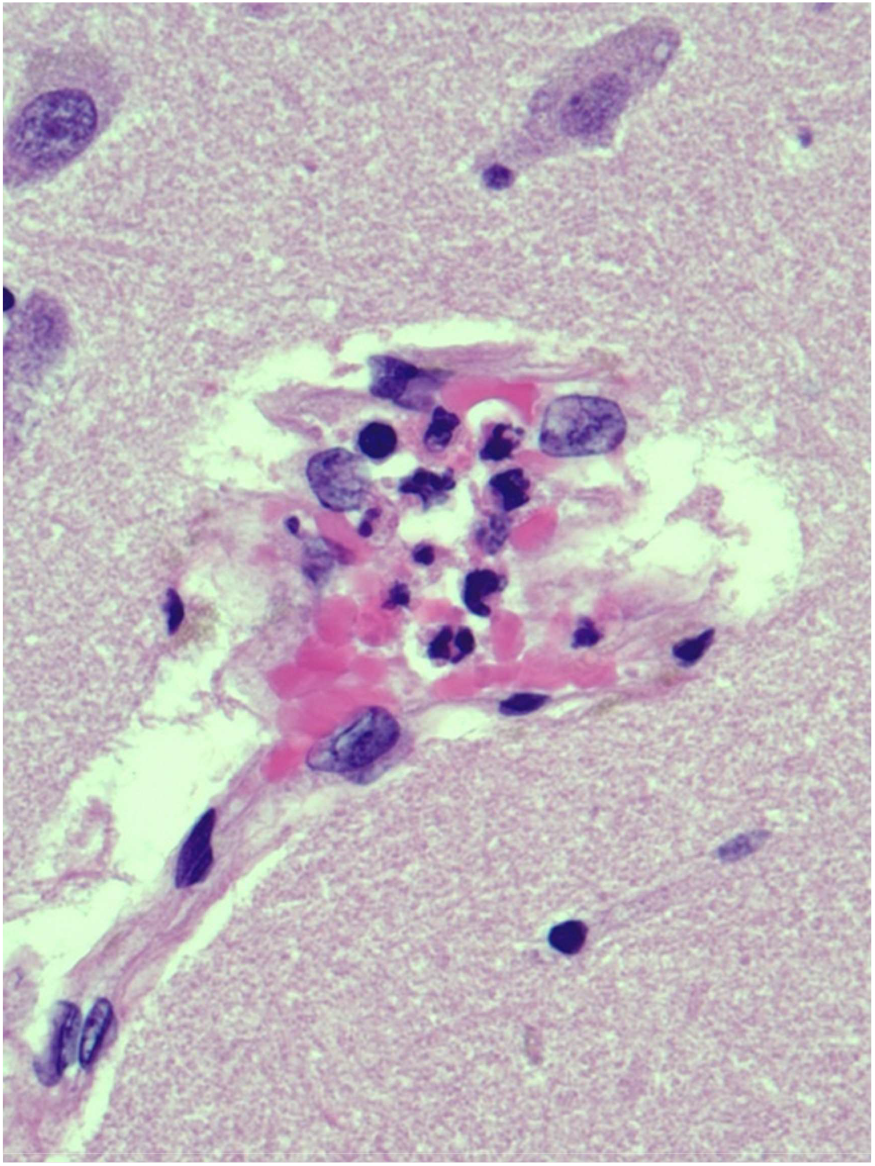
Patient 10: A microvessel in the H-1 zone of the right hippocampus has acute endotheliitis and apparent mural dehiscence, x400. H&E stain.

There was also a distinctive reactive microangiopathy in the ten COVID-19 brains. Briefly, this included both microvascular dilation and intussusceptive arborization (angiogenesis). Most of the dilated microvessels were thin-walled and distorted, and at least a few were present in almost every section examined. Microvascular mural distortion was often shaped like an ampoule where the channel seemed to bend and occasionally it was star-burst–like. Each case included a few microvessels with thin-walled, elongated, sinuous or tuft-like projections that suggested microvascular injury with stalled or incomplete mural healing. Some microvessels with mural sclerosis acquired these shapes. A frequent finding was red cell aggregation and serum proteins filling dilated microvessels, often with microthrombi present. Occlusive thrombosis was relatively infrequent.

## Discussion

These ten autopsy cases of COVID-19 vary in patient age to some extent, there is equal gender representation, and the clinical courses vary considerably in length and complexity. Most patients are hypertensive with adventitial sclerosis as evidence of microvascular wall injury. Predisposing vascular injury might leave microvessels more prone to autoimmune damage.^16^ Most patients in the cohort are African American, but given the small sample size no definite effect of race can be inferred.

It is important to point out that all ten patients have the same early stage of type 3 hypersensitivity vasculitis expressed as acute neutrophilic endotheliitis. In type 3 hypersensitivity vasculitis, PMN activation is the main vascular damage effector along with hypercytokinemia.^7^ Dehiscent microvessels identified in the cohort are consistent with type 3 hypersensitivity capillary necrosis. The occasional perivascular hemorrhage noted as well as acute perivasculitis formed of transmigrating PMNs are features of this type of autoimmune vasculitis. No fibrinoid necrosis is identified, which would have indicated a later stage of this type of vasculitis.^25,26^ Other causes of capillary necrosis might include direct SARS-CoV-2 infection or an indirect effect such as occlusive thrombosis,^27^ hypercytokinemia, or an adverse balance of the RAS.^9^ Intravascular neutrophilic aggregates and release of neutrophilic extracellular traps (NETs) have also been documented in patients with severe COVID-19,^24,28,29^ and IgG fractions isolated from patients with COVID-19 have been shown to promote NET release from neutrophils of healthy individuals.^28,29^ This mechanism has been linked to autoantibody formation as well as to vascular thrombosis.^29^

The particular significance of acute endotheliitis in this cohort appears to be that it is found as the early phase of autoimmune vasculitis in patients presenting and dying very early in the clinical course of COVID-19 as well as in those dying days, weeks, or, in one patient, months into the disease course. Multifocal acute endotheliitis suggests that its effects could cause damage anywhere in the brain and in multiple brain locations simultaneously. Damage to the central autonomic network and cardiorespiratory pacing nuclei such as the nucleus of the tractus solitarius has been raised as a concern in COVID-19. Injury to these interconnected systems, which are located in the brainstem with connections to the somatosensory cortex, insular cortex, thalamus, amygdala, and hypothalamus, might have immediate negative repercussions on respiratory and cardiac function.^3,30-33^

Onset of type 3 hypersensitivity vasculitis can follow antigenic challenge by days to weeks.^16^ Factors that might influence the timing of the onset of acute endotheliitis could include patient age, unique genome, the pace of disease due to comorbidities, or all of these factors. A recently described possible influence on the pace of autoimmunity in COVID-19 patients involves mutational alterations of interferons that are part of the disease response. Such mutations can be found in life-threatening COVID-19.^7,11^ Specific inhibitors of anaphylaxis might provide effective prophylactic treatment for terminal complement component generation in type 3 hypersensitivity vasculitis.^16^

Additional proposed influences on autoimmunity in COVID-19 include SARS-CoV-2 as an environmental trigger, physical and environmental agents, and hormonal factors. Autoimmunity might be induced by molecular mimicry and “bystander activation.” Antigen-presenting cells that are activated in the primary infection might activate pre-primed autoreactive T lymphocytes that secrete pro-inflammatory mediators that damage CNS parenchyma.^12^ Stalled or sluggish microvascular blood flow from microthrombi or from more substantial microvascular damage in COVID-19 could exacerbate functional shunting deficits in vascular beds in addition to generating damaging pro-inflammatory cytokines in the already hypoxically-challenged brain.^7^

The possibility of a central hypoventilation syndrome involving the brainstem’s central cardiopulmonary pacing network has been suggested to occur in COVID-19.^34^ Given the respiratory challenges common to COVID-19 and in this cohort, and the possibility of a central hypoventilation syndrome involving the brainstem’s central cardiopulmonary pacing network, the time of death was noted for our cohort. Temporal evidence of a failure of the switch from automatic to voluntary breathing, wherein respiratory effort ceases as normal pacing fails, was not found here.

Components of the reactive microangiopathy that include intussusceptive arborization,^33^ which has been described in the brain in COVID-19,^18^ evidence suggesting a delay in healing of injured brain microvessels, and further comments on possible effects of acute neutrophilic endotheliitis will be discussed in a later submission. Uncoupling of neurogliovascular units during microvascular injury will also be commented upon. Uncoupling of these units causes a decline in neuronal activity.^36-38^

## Data Availability

All data in the manuscript are supported by completed and signed University Medical Center-New Orleans autopsy files.

